# The impact of telemedicine on pediatric hospital capacity and resources: A study protocol for a systematic review

**DOI:** 10.1101/2024.04.25.24306362

**Authors:** JHL Wagenaar, HR Taal, IKM Reiss, MS Kleinsmann, JL Fang, S Hinrichs

## Abstract

**Background:** Telemedicine has evolved significantly, demonstrating benefits across various medical fields, including pediatrics. Hospital capacity issues and long distances to reach specialty care are common incentives to use telemedicine. However, current telemedicine research focuses primarily on its effect on clinical outcomes (efficacy) and implementation barriers, leaving capacity outcomes and effectiveness of telemedicine underexplored. This systematic review aims to fill this gap by providing an overview of telemedicine’s impact on pediatric hospital capacity.

**Methods:** The PRISMA guidelines will be followed. A systematic literature search through the Pubmed, Embase, Cochrane and Web of Science databases will be conducted. Empirical and modelling studies that evaluate the impact of telemedicine on capacity outcomes in a pediatric, neonatal or perinatal population and hospital setting will be included. Two independent researchers will perform screening, data extraction and quality assessment with the mixed methods appraisal tool. Data will be synthetized qualitatively. The primary outcome will be the impact of telemedicine on capacity outcome measures (following WHO definitions) according to each telemedicine type.

## Introduction

Telemedicine, or telehealth, is the remote delivery of health care through electronic telecommunications.(1) Over the past 70 years, it has evolved from telephonic consultations to more comprehensive telecommunications including real-time vital signs, labs, images, and visualization of the patient.(2) Initially developed to overcome long distances between caregivers and patients and provide high quality of care in rural areas, telemedicine has demonstrated benefits in diverse medical fields, such as monitoring patients with diabetes,(3) supporting cardiac rehabilitation,(4) and facilitating tele-intensive care units.(5) The capacity strain experienced in hospitals has accelerated the use-cases of telemedicine with a rapid increase observed during the COVID-19 pandemic.(6) Telemedicine has also demonstrated its potential in pediatric care,(7) where urgent capacity issues persist.(8, 9)

However, while there is potential and promise of telemedicine for reducing capacity strains, the real impact on a hospital’s capacity, once implemented, is under investigated.(10) Current research tends to focus on clinical efficacy within controlled trial settings, patient experience or implementation barriers of telemedicine, while efficiency and implementation are scarcely investigated.(11-15) Therefore, more hybrid study designs, assessing both effectiveness and implementation, are needed to estimate the impact of telemedicine in real-world settings.(16, 17)

Moreover, to the best of our knowledge, no studies have provided a comprehensive overview of pediatric telemedicine use-cases and their evidence-based impact on different aspects of hospital capacity. Such an overview is necessary for designing future telemedicine interventions, to understand the potential impact of implementation, adoption and scale-up of these interventions.

In this systematic review, we respond to this knowledge gap by creating an overview of the impact of pediatric telemedicine interventions on hospital capacity and resource use. With this overview, our study aims to advise future researchers, policymakers, and healthcare providers on effectively integrating capacity considerations into developing, implementing, and evaluating telemedicine interventions, to enhance not only clinical outcomes but also contribute to the efficient utilization of hospital resources.

## Methods

To provide an overview of impact of pediatric telemedicine on hospital capacity, a systematic review will be performed, following the PRISMA guidelines.(18) This protocol is registered in the PROSPERO database for systematic reviews (ID: CRD42024525889).

### Definitions

Telemedicine is defined as the remote delivery of health care, and grouped in 1) remote patient monitoring, 2) real-time physician to physician consultations, and 3) asynchronous (non real-time) telecommunication.(19) Digital health applications that do not deliver health care to a patient will not be included in our review. For the purposes of this study, hospital capacity is defined according to the WHO framework for health system performances as one of: 1) workforce, 2) building infrastructure and IT services, including hospital admissions, transfers and bed occupation, 3) devices and equipment, 4) pharmaceuticals.(20) Furthermore, 5) cost evaluations will also be included when related to hospital resources. Evaluation of environmental impact will not be included as hospital capacity measurement.

### Eligibility criteria

Based on the definitions as described above, the following inclusion criteria will be used:

- Population: The telemedicine intervention should be suitable for pediatric care, including neonatal care
- Intervention: The telemedicine intervention must be the primary intervention evaluated
- Comparison: there will be no restriction on comparator group.
- Outcome: Studies must measure capacity impact as an outcome
- Setting: Outcomes must be evaluated in a hospital setting
- Study design: study design must be one of the following: empirical studies (cross sectional, longitudinal, prospective, implementation science), simulation and/or modelling studies

#### Exclusion criteria

- Not available in English or Dutch
- Full text unavailable
- Non health care delivering digital health applications, e.g. electronic patient records systems, decision support tools, organizational tools, training/education purposes
- Interventions that contain solely telephonic consultations, since telephonic consultation have been standard of care for decades (21)
- Health care systems outside of the hospital, e.g. primary care and schools
- Other study types, e.g. literature reviews, commentary editorials / letters to the editors, news articles, registered trials or protocols, case studies

### Search strategy

Medline, Embase, Web of Science and Cochrane databases will be searched using the following three terms: 1) telemedicine, 2) hospital capacity, and 3) pediatrics (complete search is presented in supplemental 1).

### Screening

Title and abstracts will be screened by two independent researchers (SH, JW) supported by the Covidence program for systematic reviews. Conflicts will be resolved by discussion and when needed an extra researcher (HRT). Full text screening will be performed by both SH and JW for all articles.

### Risk of bias and data extraction

Quality assessment will be performed using the Mixed Methods Appraisal Tool.(22) Per included study the following data items will be extracted: author, year of publication, country of origin, study design, pediatric use case (targeted patients/pediatric condition, pediatric department), number of patients included, number of telemedicine consultations, number of hospitals, type of telemedicine intervention, hospital capacity measurement unit, whether capacity was the primary or secondary outcome, and the observed impact on hospital capacity.

### Data synthesis

Extracted data will be synthetized qualitatively. Data will be presented in an overview table (dummy table 1) categorized in 3 domains in order to visualize the available evidence: 1) type of telemedicine, 2) type of capacity outcome and the measured unit and 3) quality of evidence using mixed methods appraisal tool. The impact of the intervention will be summarized as an increase or decrease and will be presented per capacity outcome measure for the telemedicine types.

**Table 1.** Dummy table for data synthesis.

|  |  | Article |  | Impact on hospital capacity |  |  |  |  |  |  |  |  |  |  |  |  |  |
| --- | --- | --- | --- | --- | --- | --- | --- | --- | --- | --- | --- | --- | --- | --- | --- | --- | --- |
|  |  | <u>Information</u> | <u>Quality of evidence</u> | <u>Workforce</u> |  |  | <u>Building infrastructure and IT</u> |  |  |  |  | <u>Devices and equipment</u> |  | <u>Costs</u> |  |  |  |
|  |  | (author, year, country of origin) |  | Time invested by healthcare provider | Work-load | other | Length of stay | Transfers | Outpatient clinic visits | Hospital admissions | other | Telemedicine application | other | CEA | CMA | BIA | other |
| Type of telemedicine | Remote patient monitoring |  |  |  |  |  |  |  |  |  |  |  |  |  |  |  |  |
|  | Real-time physician to physician consultation |  |  |  |  |  |  |  |  |  |  |  |  |  |  |  |  |
|  | Asynchronous communication |  |  |  |  |  |  |  |  |  |  |  |  |  |  |  |  |

Abbreviations: CEA: cost effectiveness analysis; CMA: cost minimization analysis; BIA: budget impact analysis.

## Data Availability

All data produced in the present study are available upon reasonable request to the authors

## Funding/Acknowledgements

This research is supported by Convergence | Healthy Start, a program of the Convergence Alliance – Delft University of Technology, Erasmus University Rotterdam and Erasmus Medical Center - to improve the future of new generations.

## Supplemental 1 – search strategy

| Database searched | Platform | Years of coverage | Records | Records after duplicates removed |
| --- | --- | --- | --- | --- |
| Medline ALL | Ovid | 1946 - Present | 2750 | 2733 |
| Embase | Embase.com | 1971 - Present | 3191 | 916 |
| Web of Science Core Collection* | Web of Knowledge | 1975 - Present | 2764 | 400 |
| Cochrane Central Register of Controlled Trials | Wiley | 1992 - Present | 788 | 453 |
| **Total** |  |  | **9493** | **4502** |

*Science Citation Index Expanded (1975-present); Social Sciences Citation Index (1975-present); Arts & Humanities Citation Index (1975-present); Conference Proceedings Citation Index-Science (1990-present); Conference Proceedings Citation Index- Social Science & Humanities (1990-present); Emerging Sources Citation Index (2005-present)

No other database limits were used than those specified in the search strategies

### Medline

(exp “Telemedicine”/ OR *”Telecommunications”/ OR “Videoconferencing”/ OR (telehospital* OR tele-hospital* OR telehealth* OR tele-health* OR telemedic* OR tele-medic* OR telemonitor* OR tele-monitor* OR telecare OR tele-care OR tele-icu* OR tele-intensive-car* OR telepresence* OR tele-presence* OR tele-referral* OR telereferral* OR teleconsultat* OR teleradiol* OR tele-radiol* OR ((remot* OR electronic* OR virtual*) ADJ3 (health* OR intervention* OR consult* OR diagno* OR medicine*)) OR ((remot*) ADJ3 (monitor*)) OR ((smart*) ADJ3 (health*)) OR uhealth OR ((digital* OR remot* OR electronic* OR virtual* OR mobile) ADJ3 (distanc*) ADJ3 (practice*)) OR videoconferenc* OR video-conferenc* OR smartwatch* OR smart-watch* OR teleNICU OR tele-NICU OR tele-neo* OR teleneo*).ab,ti,kf. OR (e-health* OR ehealth* OR mhealth* OR m-health* OR ((digital*) ADJ3 (health* OR intervention* OR consult* OR diagno* OR medicine*)) OR mobile-health* OR remote-consultat*).ti.) **AND** (exp “Personnel Staffing and Scheduling”/ OR exp “Employment”/ OR exp “Nursing Staff”/ OR exp “Medical Staff”/ OR exp “Workforce”/ OR “Length of Stay”/ OR exp “Hospital Bed Capacity”/ OR (((length*) ADJ3 (stay*)) OR ((hospital* OR patient* OR intrahospital* OR intra-hospital* OR interhospital* OR inter-hospital* OR NICU OR neonatal-intensive-care-unit* OR child* OR pediatr* OR paediatr*) ADJ3 (admission* OR admittan* OR admitting OR discharg* OR bed OR beds OR transfer* OR transport*)) OR waiting-list* OR ((discharg* OR postdischarg*) ADJ3 (plan* OR process*)) OR ((bed OR beds) ADJ3 (utiliz* OR utilis*)) OR ((effect* OR utili*) ADJ3 (cost*)) OR resourc*-generat* OR workforce* OR staff* OR work-forc* OR personnel* OR capacit* OR implement* OR hybrid*-design OR efficien*).ab,ti,kf.) **AND** (exp *”Pediatrics”/ OR exp *”Child”/ OR exp *”Infant”/ OR *”Adolescent”/ OR (pediatr* OR paediatr* OR peadiatr* OR neonat* OR newborn* OR child OR children OR boy OR boys OR girl OR girls OR infant* OR toddler* OR teenager* OR adolescen* OR teen OR teens OR teleNICU OR tele-NICU OR tele-neo* OR teleneo*).ti.) **NOT** (news OR congres* OR abstract* OR book* OR chapter* OR dissertation abstract*).pt.

### Embase

(‘telemedicine’/de OR ‘telediagnosis’/de OR ‘teleconsultation’/de OR ‘telesurgery’/de OR ‘telecare’/exp OR ‘telehealth’/exp OR ‘telemonitoring’/exp OR ‘telecommunication’/mj/de OR ‘digital health’/exp/mj OR ‘digital health technology’/exp OR ‘digital health intervention’/exp OR ‘electronic consultation’/exp OR ‘mhealth’/exp OR ‘mobile health’/exp/mj OR ‘videoconferencing’/de OR (telehospital* OR tele-hospital* OR telehealth* OR tele-health* OR telemedic* OR tele-medic* OR telemonitor* OR tele-monitor* OR telecare OR tele-care OR tele-icu* OR tele-intensive-car* OR telepresence* OR tele-presence* OR tele-referral* OR telereferral* OR teleconsultat* OR teleradiol* OR tele-radiol* OR ((remot* OR electronic* OR virtual*) NEAR/3 (health* OR intervention* OR consult* OR diagno* OR medicine*)) OR ((remot*) NEAR/3 (monitor*)) OR ((smart*) NEAR/3 (health*)) OR uhealth OR ((digital* OR remot* OR electronic* OR virtual* OR mobile) NEAR/3 (distanc*) NEAR/3 (practice*)) OR videoconferenc* OR video-conferenc* OR smartwatch* OR smart-watch* OR teleNICU OR tele-NICU OR tele-neo* OR teleneo*):ab,ti,kw OR (e-health* OR ehealth* OR mhealth* OR m-health* OR ((digital*) NEAR/3 (health* OR intervention* OR consult* OR diagno* OR medicine*)) OR mobile-health* OR remote-consultat*):ti) **AND** (‘personnel management’/exp OR ‘employment’/exp OR ‘nursing staff’/exp OR ‘medical staff’/exp OR ‘workforce’/exp OR ‘length of stay’/exp OR ‘hospital transport system’/exp OR ‘hospital admission’/exp OR ‘hospital discharge’/exp OR ‘hospital bed capacity’/exp OR ‘hospital bed’/exp OR ‘hospital bed utilization’/exp OR ‘patient transport’/exp OR (((length*) NEAR/3 (stay*)) OR ((hospital* OR patient* OR intrahospital* OR intra-hospital* OR interhospital* OR inter-hospital* OR NICU OR neonatal-intensive-care-unit* OR child* OR pediatr* OR paediatr*) NEAR/3 (admission* OR admittan* OR admitting OR discharg* OR bed OR beds OR transfer* OR transport*)) OR waiting-list* OR ((discharg* OR postdischarg*) NEAR/3 (plan* OR process*)) OR ((bed OR beds) NEAR/3 (utiliz* OR utilis* OR capacit*)) OR ((effect* OR utili*) NEAR/3 (cost*)) OR resourc*-generat* OR workforce* OR staff* OR work-forc* OR personnel* OR capacit* OR implement* OR hybrid*-design OR efficien*):ab,ti,kw) **AND** (‘pediatrics’/exp/mj OR ‘child’/exp/mj OR ‘adolescent’/exp/mj OR (pediatr* OR paediatr* OR peadiatr* OR neonat* OR newborn* OR child OR children OR boy OR boys OR girl OR girls OR infant* OR toddler* OR teenager* OR adolescen* OR teen OR teens OR teleNICU OR tele-NICU OR tele-neo* OR teleneo*):ti) **NOT** ([Conference Abstract]/lim OR [preprint]/lim)

### Web of Science

(TS=( telehospital* OR tele-hospital* OR telehealth* OR tele-health* OR telemedic* OR tele-medic* OR telemonitor* OR tele-monitor* OR telecare OR tele-care OR tele-icu* OR tele-intensive-car* OR telepresence* OR tele-presence* OR tele-referral* OR telereferral* OR teleconsultat* OR teleradiol* OR tele-radiol* OR ((remot* OR electronic* OR virtual*) NEAR/2 (health* OR intervention* OR consult* OR diagno* OR medicine*)) OR ((remot*) NEAR/2 (monitor*)) OR ((smart*) NEAR/2 (health*)) OR uhealth OR ((digital* OR remot* OR electronic* OR virtual* OR mobile) NEAR/2 (distanc*) NEAR/2 (practice*)) OR videoconferenc* OR video-conferenc* OR smartwatch* OR smart-watch* OR teleNICU OR tele-NICU OR tele-neo* OR teleneo*) OR TI=( e-health* OR ehealth* OR mhealth* OR m-health* OR ((digital*) NEAR/2 (health* OR intervention* OR consult* OR diagno* OR medicine*)) OR mobile-health* OR remote-consultat*)) **AND** TS=(((length*) NEAR/2 (stay*)) OR ((hospital* OR patient* OR intrahospital* OR intra-hospital* OR interhospital* OR inter-hospital* OR NICU OR neonatal-intensive-care-unit* OR child* OR pediatr* OR paediatr*) NEAR/2 (admission* OR admittan* OR admitting OR discharg* OR bed OR beds OR transfer* OR transport*)) OR waiting-list* OR ((discharg* OR postdischarg*) NEAR/2 (plan* OR process*)) OR ((bed OR beds) NEAR/2 (utiliz* OR utilis* OR capacit*)) OR ((effect* OR utili*) NEAR/2 (cost*)) OR resourc*-generat* OR workforce* OR staff* OR work-forc* OR personnel* OR capacit* OR implement* OR hybrid*-design OR efficien*) **AND** TI=(pediatr* OR paediatr* OR peadiatr* OR neonat* OR newborn* OR child OR children OR boy OR boys OR girl OR girls OR infant* OR toddler* OR teenager* OR adolescen* OR teen OR teens OR teleNICU OR tele-NICU OR tele-neo* OR teleneo*) **NOT** DT=(Meeting Abstract OR Meeting Summary)

### Cochrane CENTRAL

((telehospital* OR tele NEXT/1 hospital* OR telehealth* OR tele NEXT/1 health* OR telemedic* OR tele NEXT/1 medic* OR telemonitor* OR tele NEXT/1 monitor* OR telecare OR tele NEXT/1 care OR tele NEXT/1 icu* OR tele NEXT/1 intensive NEXT/1 car* OR telepresence* OR tele NEXT/1 presence* OR tele NEXT/1 referral* OR telereferral* OR teleconsultat* OR teleradiol* OR tele NEXT/1 radiol* OR ((remot* OR electronic* OR virtual*) NEAR/3 (health* OR intervention* OR consult* OR diagno* OR medicine*)) OR ((remot*) NEAR/3 (monitor*)) OR ((smart*) NEAR/3 (health*)) OR uhealth OR ((digital* OR remot* OR electronic* OR virtual* OR mobile) NEAR/3 (distanc*) NEAR/3 (practice*)) OR videoconferenc* OR video NEXT/1 conferenc* OR smartwatch* OR smart NEXT/1 watch* OR teleNICU OR tele NEXT/1 NICU OR tele NEXT/1 neo* OR teleneo*):ab,ti,kw OR (e NEXT/1 health* OR ehealth* OR mhealth* OR m NEXT/1 health* OR ((digital*) NEAR/3 (health* OR intervention* OR consult* OR diagno* OR medicine*)) OR mobile NEXT/1 health* OR remote NEXT/1 consultat*):ti) **AND** ((((length*) NEAR/3 (stay*)) OR ((hospital* OR patient* OR intrahospital* OR intra NEXT/1 hospital* OR interhospital* OR inter NEXT/1 hospital* OR NICU OR neonatal NEXT/1 intensive NEXT/1 care NEXT/1 unit* OR child* OR pediatr* OR paediatr*) NEAR/3 (admission* OR admittan* OR admitting OR discharg* OR bed OR beds OR transfer* OR transport*)) OR waiting NEXT/1 list* OR ((discharg* OR postdischarg*) NEAR/3 (plan* OR process*)) OR ((bed OR beds) NEAR/3 (utiliz* OR utilis* OR capacit*)) OR ((effect* OR utili*) NEAR/3 (cost*)) OR resourc* NEXT/1 generat* OR workforce* OR staff* OR Work NEXT/1 forc* OR personnel* OR capacit* OR implement* OR hybrid* NEXT/1 design OR efficien*):ab,ti,kw) **AND** ((pediatr* OR paediatr* OR peadiatr* OR neonat* OR newborn* OR child OR children OR boy OR boys OR girl OR girls OR infant* OR toddler* OR teenager* OR adolescen* OR teen OR teens OR teleNICU OR tele NEXT/1 NICU OR tele NEXT/1 neo* OR teleneo*):ti)

